# Stochastic Transmission in Epidemiological Models

**DOI:** 10.1101/2023.01.15.23284574

**Authors:** Vinicius V.L. Albani, Jorge P. Zubelli

## Abstract

Recent empirical evidence suggests that the transmission coefficient in susceptible-exposed-infected-removed-like (SEIR-like) models evolves with time, presenting random patterns, and some stylized facts, such as mean-reversion and jumps. To address such observations we propose the use of jump-diffusion stochastic processes to parameterize the transmission coefficient in an SEIR-like model that accounts for death and time-dependent parameters. We provide a detailed theoretical analysis of the proposed model proving the existence and uniqueness of solutions as well as studying its asymptotic behavior. We also compare the proposed model with some variations possibly including jumps. The forecast performance of the considered models, using reported COVID-19 infections from New York City, is then tested in different scenarios, including major outbreaks. The proposed jump-diffusion model presented remarkably accurate out-of-sample predictions, even during larger forecasted periods.

## 1 Introduction

Since the beginning of the Coronavirus Disease 2019 (COVID-19) pandemic, numerous models were proposed to describe, calibrate and forecast the SARS-CoV-2 virus spread dynamics, as well as, the disease evolution in populations. Some models are agent-based [42] or network-based [1], other are generalizations of the classical susceptible-infected-removed (SIR) or susceptible-exposed-infected-removed (SEIR) models [3, 7, 10, 17, 23, 32]. Some models are based on partial-differential equations [35], statistical modeling [55], or yet, neural networks [47]. Other works use multi-scale approaches, which consider, for example, the interaction among individuals and between the virus and cells [15, 16].

One of the main difficulties faced in modeling is converting data into useful information, such as accurate forecasts to help sanitary authorities to implement contention or mitigation measures. These authorities must also appropriately adjust the intensity of lockdowns and movement restrictions to minimize the impact on the labor market and the economy in general. In fact, during 2020, many countries implemented socioeconomic measures to alleviate the impact of lockdowns, furnishing income to those that lost their jobs [2, 4], which could have helped the stay-at-home policies.

Another difficulty faced in modeling is balancing model parsimony and prediction accuracy [17]. A highly sophisticated model can overfit the data producing accurate in-sample predictions but misleading forecasts. In other words, the model must be as simple as possible, but not too simple.

A series of recent articles used SIR or SEIR-like models to describe the COVID-19 dynamics accounting for time-dependent transmission parameters that were estimated from reported infections [3, 6, 5, 7, 21, 22, 11]. These estimations provided empirical evidence for the time-dependency and the stochastic nature of the SARS-CoV-2 transmission [3]. For other diseases, such as measles, such time variation was already investigated [19].

It is well-known that, in SIR and SEIR-like models, the transmission parameter is a function of the average number of contacts someone has in a time period and the probability of each of these contacts with an infected being will cause transmission [41]. Intuitively, there is no reason for such values to stay static over time. Moreover, the probability of transmission deeply depends on the pathogen evolution, which is highly uncertain. Thus, it seems natural to parameterize the transmission coefficient as a stochastic process, which is a random process that has a distribution that evolves with time. In [3, 45], the Cox-Ingersoll-Ross (CIR) model [26], from Mathematical Finance, was proposed to describe the transmission parameter dynamics in SEIR-like models. In fact, in [3], the resulting SEIR-like model produced accurate forecasts with different time horizons using datasets from different places. The rationale behind the use of this model is its mean-reversion which seems to be one of the main characteristics of the transmission parameter values estimated from data.

The use of stochastic models to describe the evolution of epidemics in populations has a long-standing tradition, and dates back at least, to the 1950s and 1960s [14, 13, 56]. For example, in [54], the studied model was based on a discrete-time Markov chain. More recently, some authors proposed stochastic versions of the susceptible-infected (SI) model [39, 48] studying its asymptotic behavior, based on the so-called stochastic perturbation of parameters. Some stochastic SI- or SEIR-like models were also proposed in [28, 30, 34, 53, 57]. In fact, the reference [53] proposes a stochastic SIR model with jumps. A review of stochastic epidemiological models can be found in [20].

The main difference between our approach with these previous works is the direct modeling of the transmission parameter as a stochastic process. This allows us to define a structure for the transmission parameter accounting for observed stylized facts, such as mean-reversion and jumps. Moreover, by assuming that the only source of uncertainty comes from transmission, we avoid adding unnecessary stochastic terms to the epidemiological dynamics, keeping the model parsimonious and avoiding dealing with cross-correlations, that are difficult to estimate from data. We shall see that considering only the transmission parameter as a stochastic process, is sufficient to provide adherence to data and accurate forecasts.

In summary, the article’s contributions are as follows, a general jump-diffusion mean-reverting model is proposed to describe the dynamics of the transmission parameter in an SEIR-like epidemiological model. The resulting model is then analyzed theoretically, i.e., results on the existence and uniqueness of solutions are proved. Concerning the asymptotic behavior of the model, we consider the versions with and without jumps. In both cases, the existence of invariant or asymptotic measures is presented using recent results on the Ergodic properties of stochastic processes. For the jumpless version, a closed-formula for the invariant measure is achieved. For the model with jumps, the invariant measure is given by the solution of an integro-differential equation that is solved numerically. We also consider in the analysis and in the numerical examples the classical CIR model, and its version with jumps. We test the forecast performance of the presented models using COVID-19 reported infections from New York City (NYC), paying special attention to two major outbreaks, namely, the second outbreak of 2020 and the outbreak caused by the omicron variant at the end of 2021.

The article is organized as follows, Section 2 introduces the epidemiological model, as well as existence and uniqueness results are presented. Section 2 also recalls the asymptotic properties of the deterministic SEIR model. The asymptotic analysis of different versions of the model to the transmission parameter is developed in Section 3. Section 4 presents some numerical examples evaluating predictions given by the epidemiological model considering different versions of the dynamics of the transmission parameter using data from NYC. It also presents an example that accounts for reinfection and loss of immunity. Concluding remarks and a further discussion about the modeling are drawn in Section 5. Appendix A.1 presents the details of the implementation of the numerical examples in the text.

## 2 The Epidemiological Model

### 2.1 The General Setting

Consider the filtered probability space defined by (Ω, ℱ, {ℱ_*t*_}_*t*≥0_, ℙ), where Ω is the sample space, ℱ is a *σ*-algebra defined on Ω, {ℱ_*t*_}_*t*≥0_ is a filtration, and ℙ a probability measure defined on (Ω, ℱ) [40, 51]. The proposed SEIR-type model accounts for susceptible (S), exposed (E), infected (I), recovered (R), and deceased (D) compartments. Individuals progress from one compartment to another accordingly to the system of differential equations below:

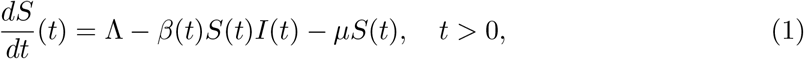

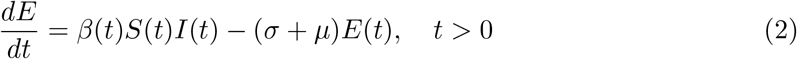

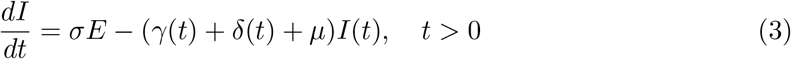

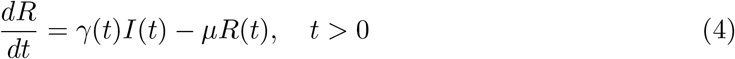

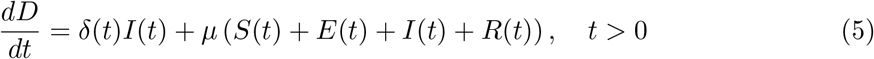

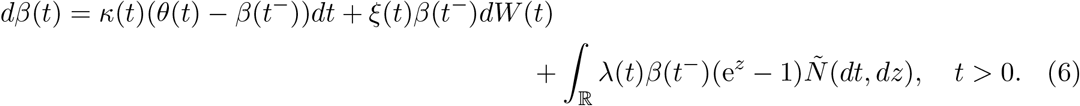

The time-dependent transmission parameter *β*(*t*) is a jump-diffusion, satisfying the stochastic differential equation (SDE) [52] in Eq. (6), where *W* (*t*) is a Brownian motion [40] and *Ñ* (*dt, dz*) is a compensated version of the Poisson random measure [52, 24]. In other words, there exists a *σ*-finite measure *ν*(*dz*) such that *Ñ* (*dt, dz*) = *N* (*dt, dz*) − *ν*(*dz*)*dt*, and *N* (*dt, dz*) is the Poisson random measure. The transmission has two main features, it is mean-reverting and non-negative. The first two parts of the right-hand side (RHS) of the SDE in Eq. (6) is the diffusive part, whereas the third part is the jump part. The compensator measure *ν*(*dz*) is the Lévy measure of the process, which is the so-called jump-size distribution, and the Poisson measure *N* (*dt, dz*) states the time-frequency of jumps with certain sizes. In general, small jumps can be modeled as diffusion, however, the transmission can suffer dramatic regime changes that are represented by large jumps in the time series of estimated values for *β*(*t*). Such large jumps are observed, for example, when a more transmissible variant arises. Thus, allowing the process that defines the transmission rate jump can be more realistic.

For simplicity, we assume that only the transmission parameter has stochastic dynamics since our aim is to understand how the stochastic perturbation of the transmission parameter interferes with the qualitative behavior of SEIR-type models.

It is worth noticing that, if *ω* ∈ Ω is fixed, *β*(*t*) = *β*(*t, ω*) as a function of *t* is continuous by parts. More precisely it is a right-continuous with left limit (càdlàg), this is why we use the superscript *minus* sign (−) in the SDE in Eq. (6). Thus, for almost every *ω*, we can analyze the system of ordinary differential equations in Eqs. (2)–(5) using usual techniques.

### 2.2 Existence and Uniqueness of Solutions

Firstly, we must state that there exists a unique non-negative solution of the SDE in Eq. (6).

#### Lemma 1.

*Assume that the coefficients κ*(*t*), *θ*(*t*), *ξ*(*t*), *and λ*(*t*) *are non-negative and bounded functions. In particular*, 0 ≤ *λ*(*t*) ≤ 1. *Assume also that the jump-size distribution satisfies:*

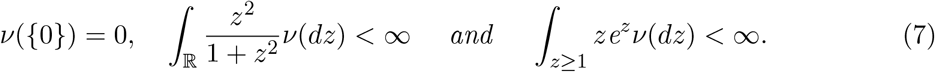

*There exists a unique non-negative and càdlàg process β*(*t*) *satisfying the SDE in* *Eq*. (6) *for any deterministic initial condition β*(0) = *β*_0_ *>* 0.

*Proof*. The first two conditions in Eq. (7) state that *ν* is a Lévy measure. The third condition states that the integral part in the SDE in Eq. (6) is finite. The existence and uniqueness of a càdlàg process *β*(*t*) satisfying the SDE in Eq. (6), given a deterministic initial condition *β*(0) = *β*_0_ *>* 0, follows by [52, Theorem 1.19], since the parameters *α*(*t, x*) = *κ*(*t*)(*θ*(*t*) − *x*), *σ*(*t, x*) = *ξ*(*t*)*x*, and *γ*(*t, x, z*) = *λ*(*t*)*x*(e^*z*^ −1) satisfy the linear growth and Lipschitz continuity on the variable *x* conditions. Moreover, the expected value 𝔼[*β*(*t*)^2^] is finite for every *t* ≥ 0.

The non-negativity follows since *β*(*t*) has the formula

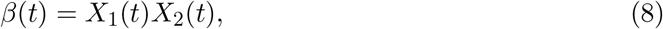

where

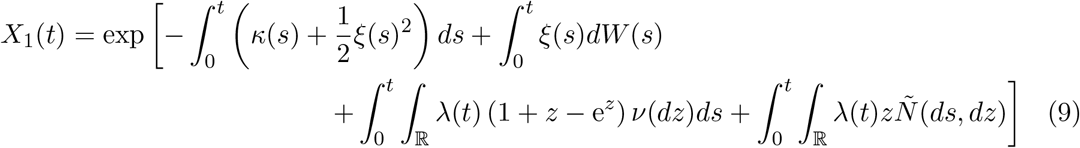

and

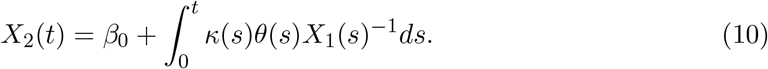

Since *X*_1_(*t*) ≥ 0 almost surely (a.s.), it follows that *x*_2_(*t*) is also non-negative, since *β*_0_, *κ*(*t*), and *θ*(*t*) are non-negative. Thus, *β*(*t*) is a.s. non-negative. □

#### Remark 1.

*To obtain the formulas in* *Eqs*. (8)–(9) *for the solution of β we adapted techniques to find solutions for a general linear SDE from [29, Example 2] to the context of jump-diffusions and applied the multi-dimensional version of the Itô*’*s lemma to jump-diffusions in [52, Theorem 1*.*16]. The formula in* *Eq*. (10) *is the version with time-dependent parameters of the solution of the geometric Lévy process obtained in [52, Example 1.15] and γ*(*t, z*) = *λ*(*t*)(*e*^*z*^ − 1).

We now pass to the existence and uniqueness of the solution of the system in Eqs. (2)–(5).

#### Lemma 2.

*Let σ be a positive constant and γ*(*t*) *and δ*(*t*) *be bounded and non-negative continuous functions. Then, the system of ordinary differential equations (ODE) in* *Eqs*. (2)– (5) *has a unique solution almost surely, given appropriate initial conditions*.

*Proof*. The almost surely part comes from the fact that the SDE in Eq. (6) is satisfied almost surely by the càdlàg process *β*. Since for each *ω* ∈ Ω, *β*(*t*) = *β*(*t, ω*) is continuous by parts function and the other parameters in the ODE system in Eqs. (2)–(5) are continuous and bounded functions, existence and uniqueness follow by Theorems 1–2 in [31]. □

A simple adaptation of the calculations in [46, Section 2] shows that, for a positive initial condition, the solution for the system in Eqs. (2)–(5), with *β*(*t*) given, is non-negative. Moreover, for any initial condition such that (*S*(0), *E*(0), *I*(0), *R*(0)) is inside the region

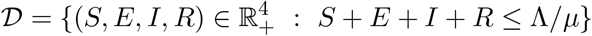

the corresponding solution remains in 𝒟, i.e., 𝒟 is positively-invariant.

We expect that, when *t* → ∞, the coefficients in the epidemiological model converge to constant values. In the analysis of the asymptotic behavior of the model in Eqs. (2)–(6), we shall assume that all coefficients, including those in the definition of *β*(*t*), do not depend on time *t*. We shall also need the definitions below.

#### Definition 1.

*Let f* (*x*) *be a bounded function with bounded first and second derivatives, then, the infinitesimal generator of the stochastic process in* *Eq*. (6) *is the operator defined as*

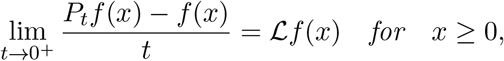

*where P*_*t*_*f* (*x*) = 𝔼 [*f* (*β*(*t*)) |*β*(0) = *x*] *is the semi-group associated with β*(*t*).

#### Definition 2.

*An invariant measure μ associated with the stochastic process β*(*t*) *in* *Eq*. (6) *is a Borel probability measure defined in* ℝ_+_ *such that*

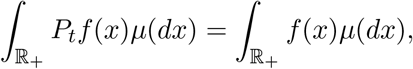

*i*.*e*.,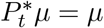.

Notice that, Definitions 1–2 can be extended to general stochastic processes. See, for example, [8, 40, 49].

### 2.3 Asymptotic Properties of the Deterministic Model

In this section, we assume that the model in Eqs. (2)–(5) has constant parameters, i.e., *β*(*t*) = *β, γ*(*t*) = *γ*, and *δ*(*t*) = *δ*. Since the asymptotic analysis of this model is well-known, we just recall some results that are relevant to the analysis that follows. We refer the readers to the works [46, 44] for more details.

In this case, the time-dependent reproduction number, obtained through the next-generation matrix method [27], is given by

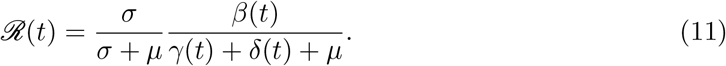

If we assume that *β* and *γ* are strictly positive constants, the resulting ODE system has two steady-state equilibrium points, namely, one disease-free equilibrium point (DFE), and one endemic equilibrium point (EEP). By steady state points, we mean limit points to the orbits of the model when *t* → ∞. In this case, the time derivatives in Eqs. (2)–(5) vanish, and the resulting algebraic system is solved for (*S*e, *E*e, *I*e, *R*e), in the case of the EEP, and for (*S*_*f*_, 0, 0, 0) in the case of DFE.

The DFE is given by *S*_*f*_ = Λ*/μ* and the EEP is given by,

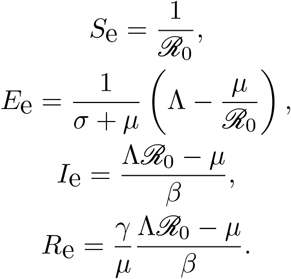

Based on results from [46, 44] it is possible to find an appropriate Lyapunov function and to verify that both equilibrium points are dynamically stable. Intuitively, dynamical stability means that, under certain conditions, the system’s solution will necessarily converge to these equilibrium points, even under small perturbations in the initial conditions.

## 3 Asymptotic Analysis of the Transmission Parameter

In this section, we study the asymptotic properties of the transmission parameter providing sufficient conditions for the existence and uniqueness of the associated invariant distribution. This analysis uses deep results in the Ergodic Theory of stochastic processes. In general, we must show that the stochastic processes satisfy a series of conditions that are related to, for example, the strong Markov property. For more details, see the notes by Martin Hairer [36] and the literature review in [9]. We start by considering the transmission without jumps, since, in this case, we can find a closed formula for the asymptotic density. This is of particular interest as we can easily evaluate the probability of the epidemiological model to reach the disease-free and the endemic equilibrium points based on the estimated values of the transmission parameter. Then, we analyze the case with jumps, providing similar results. However, in this case, the associated forward Kolmogorov equation has an integral part, which makes it difficult to find a closed formula for its solution.

### 3.1 The Transmission Parameter without Jumps

To study the asymptotic behaviour of the transmission parameter, we shall assume that its coefficients are constant, i.e., we set *κ*(*t*) = *κ, θ*(*t*) = *θ*, and *ξ*(*t*) = *ξ*. Moreover, for simplicity, in this section we also assume that the model has no jumps, i.e., we set *λ*(*t*) = 0. Thus

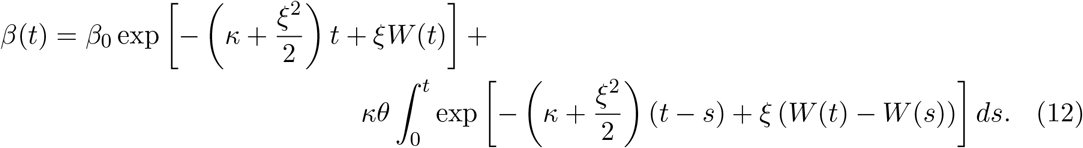

In addition, the SDE for *β*(*t*) now reads

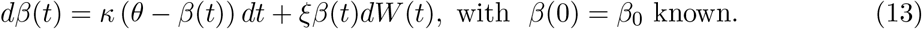

In this case, by applying Itô’s formula, Fubini’s theorem, and simple calculations to obtain the solution of linear ordinary differential equations, it follows that,

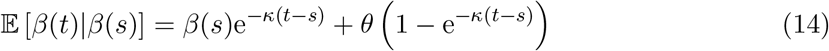

and

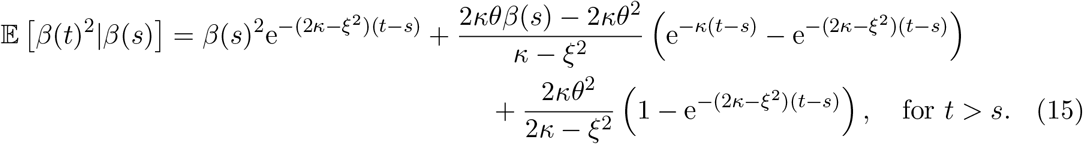

For the equality above to make sense, we must have *κ* ≠ *ξ*^2^ and 2*κ* ≠ *ξ*^2^. Based on such calculations, it is easy to evaluate 𝕍ar[*β*(*t*)] = 𝔼 *β*(*t*)^2^ − 𝔼 [*β*(*t*)]. We are interested in the behavior of *β*(*t*) when *t* → +∞. Since *κ >* 0, it follows that

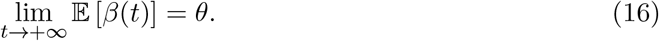

If, *κ > ξ*^2^*/*2 and *κ* ≠ *ξ*^2^,

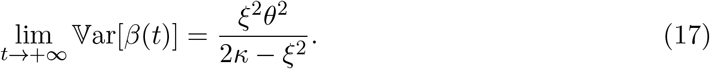

On the other hand, if *κ < ξ*^2^*/*2, and *β*_0_ satisfies

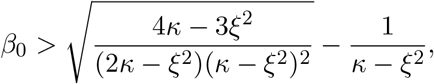

then, lim_*t*→+∞_ 𝕍ar[*β*(*t*)] = +∞.

In what follows, we assume that

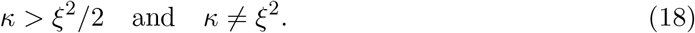

Using the estimates for 𝔼 [*β*(*t*)] and 𝕍ar[*β*(*t*)] in Eqs (14)–(15), it is possible to evaluate estimates based on the Markov and Chebychev inequalities, which are given, respectively, as follows:

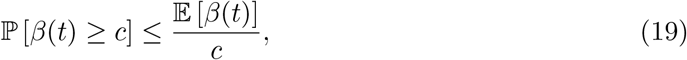

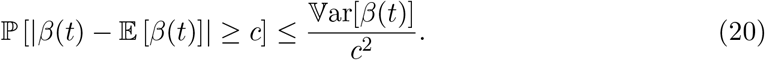

Notice that, when *t* → +∞, *W* (*t*)*/t* converges almost surely to zero. Thus, the first part of the right-hand-side (RHS) in Eq. (12) converges to zero, which means that only the integral part is relevant to the asymptotic behavior of *β*(*t*).

Another simple calculation leads us to the candidate of steady-state or invariant distribution of the stochastic process *β*(*t*). In other, words, we consider the solution of the steady-state forward Kolmogorov partial differential equation (PDE) associated with the infinitesimal generator of the process *β*(*t*). See [40].

Assume that *β*_0_ *>* 0 and *β*_0_ ≠ *θ*, then, *κ*(*θ* − *β*_0_) ≠ 0, moreover, *κ*(*θ* − *β*_0_)*ξβ*_0_ ≠ 0. In other words, *β*(*t*) satisfies the Hörmander conditions and it admits an infinitely differentiable density [49]. Such density is a solution for the forward Kolmogorov PDE [40]. Since we are interested in the asymptotic behavior of *β*(*t*), we must find the invariant measure of *β*(*t*), that, in principle, must solve the following steady-state forward Kolmogorov PDE,

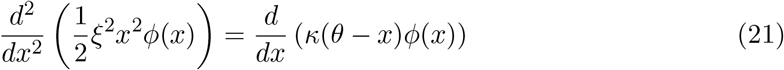

If we assume that *ϕ*(0) = 0, integrating from 0 to *x* we have

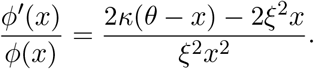

It follows that

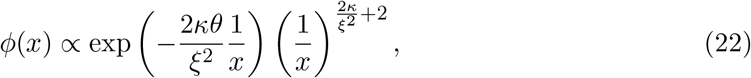

which means that 1*/β*_∞_ = lim_*t*→+∞_ 1*/β*(*t*) is Gamma-distributed, if the process *β*(*t*) admits a unique steady-state or invariant distribution. Thus, we can state the following proposition:

#### Proposition 1.

*If the hypotheses in* *Eq*. (18) *hold, then the stochastic process β*(*t*) *in* *Eq*. (12) *admits a unique invariant distribution, with the corresponding random variable denoted by β*_∞_. *In addition*, 1*/β*_∞_ *follows the Gamma distribution, i*.*e*.,

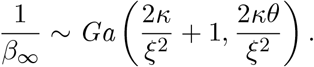

*Proof*. By assuming that the density of *β*_∞_ is *ϕ*,

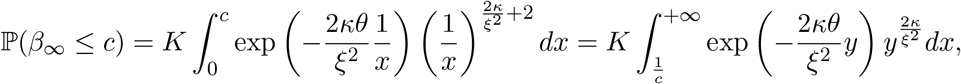

where

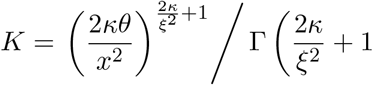

and Γ denotes the Gamma function. By applying the change of variable *y* = 1*/x* and the well-known property of Gamma functions Γ(*x* +1) = *x*Γ(*x*), it follows that the mean value and the variance of *β*_∞_ evaluated using the density *ϕ* coincide with the estimates in Eqs. (16)–(17), respectively.

The existence and uniqueness of the asymptotic or invariant measure for the process in Eq. (12) follows by the existence of a so-called strong Lyapunov function *w*(*x*), which, in the present case, can be defined as *w*(*x*) = *x*. Such strong Lyapunov function is related to the infinitesimal operator

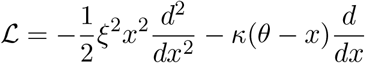

associated with *β*(*t*) in Eq. (12) and satisfies the properties, lim_*x*→+∞_ *w*(*x*) = +∞ and lim_*x*→+∞_ ℒ*w*(*x*) = +∞, since ℒ*w*(*x*) = *κ*(*x* − *θ*). Then, the hypotheses of Theorem 3.11 in [18] are satisfied and the existence and uniqueness of the invariant measure hold. Alternatively, using the same strong Lyapunov function, it is possible to use the conditions proposed in [36], as in the proof of Proposition 2 in [58]. By, the uniqueness result, the invariant distribution is given by the density in Eq. (22). □

In other words, we are considering the convergence in law, i.e., the convergence of the probability distribution of *β*(*t*) to some asymptotic distribution. In fact, there are convergence rates results when considering the semigroup associated to the distribution of *β*(*t*) and the corresponding invariant distribution. See [36]. Based on the steady-state distribution for *β*(*t*), it is possible to find the asymptotic distribution of the time-dependent reproduction number ℛ (*t*).

#### Definition 3.

*We define the steady-state effective reproduction number as*

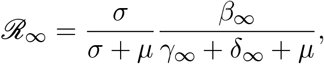

*where β*_∞_ *is the random variable associated to β*(*t*), *as well as γ*_∞_ *and δ*_∞_ *are the limit values of γ*(*t*) *and δ*(*t*), *respectively*.

Notice that, ℛ *R*_∞_ is the limit in law of ℛ (*t*) when *t* → ∞. Thus,

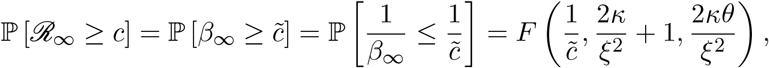

where 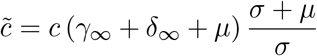 and *F* represents the Gamma cumulative distribution with the parameters 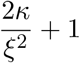 and 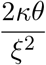.

#### Corollary 1.

*Under the assumptions of Proposition 1, the probability of the steady-state effective reproduction number to be larger than one is*

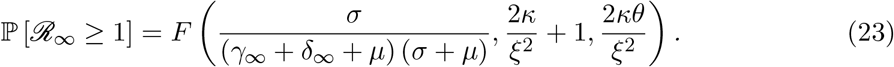

Notice that, intuitively, when *t* → +∞, we can assume that *β*(*t*) ≈ *β*_∞_, i.e., the transmission parameter is random but constant in time. In this case, it makes sense to evaluate the probability of the model in Eqs. (2)–(5) will reach the DFE and the EEP, depending on the values of the parameters of *β*, i.e., *κ, θ*, and *ξ*.

#### Example 1.

*In this example we evaluate the probability of reaching the EEP, i*.*e*., *when* ℙ (ℛ _∞_ ≥ 1), *for different values of κ, θ, and ξ and using the estimate in* *Eq*. (23). *We set μ* = 0, *γ*_∞_ = 1*/*12 *days*^−1^, *and δ*_∞_ = 1*/*14 *days*^−1^. *The parameter κ ranges from* 1 *to* 2, *θ assumes the values* 0.01, 0.05, 0.10, 0.20, 0.30, *and* 0.40, *and ξ is set to* 0.20, 0.60, *and* 1.00. *We aim to evaluate the impact of the volatility level, the mean-reverting speed, and the mean value on the probability of reaching an endemic equilibrium* ℛ _∞_ ≥ 1. *In this example*, ℙ (ℛ _∞_ ≥ 1) = ℙ (*β*_∞_ ≥ 0.155).

*When κ assumes lower values, even when θ is substantially larger than* 0.155, *we can see that the volatility ξ is more relevant, since we observe probability values considerably smaller than one. As the value of κ increases, θ becomes more relevant. This is expected as the variance of β*_∞_ *decreases when κ increases. Notice that, for simplicity, we included also cases when the first condition in* *Eq*. (18) *is violated. They are displayed on the left-hand side of the vertical solid line in the three panels of Figure 1*.

**Figure 1:**
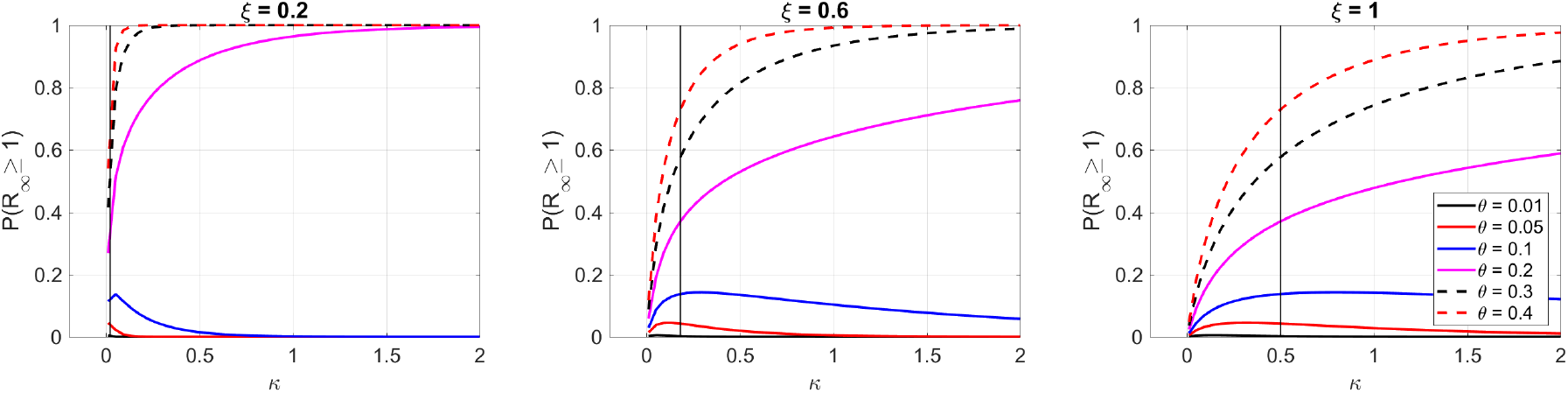
Probability of ℛ _∞_ ≥ 1 with different values for *κ, θ*, and *ξ*. The vertical line indicates when *κ* = *ξ*^2^*/*2. The probability values for *κ* ≤ *ξ*^2^*/*2 are included for completeness.

#### Remark 2.

*An important mean-reverting stochastic model that is closely related to the one in* *Eq*. (12) *is the Cox-Ingersoll-Ross (CIR) model [26] which was originally introduced to describe bond prices. This model was used in SEIR-like models to describe the dynamics of the transmission parameters in recent works [3, 45]. In this case, the parameter β satisfies the SDE below:*

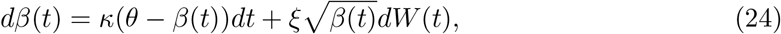

*with β*(0) = *β*_0_ *known. Unfortunately, this model does not have an explicit expression for its solution. Many of its features and asymptotic properties are well-known and well-understood. The model has a unique stable distribution [58]. If we denote the random variable associated to this distribution by β*_∞_, *it follows that*

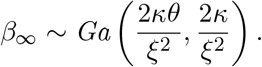

*To find the stable distribution, it is only necessary to repeat the steps used in the case of β given in* *Eq*. (12). *Notice that, in the present case*,

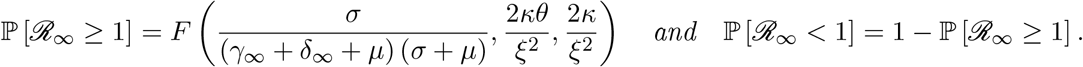

### 3.2 The Transmission Parameter with Jumps

We will now analyze the existence, uniqueness and some properties of the invariant or steadystate distribution associated to the process *β*(*t*) satisfying the SDE in Eq. (6) with the constant parameters *κ*(*t*) = *κ, θ*(*t*) = *θ, ξ*(*t*) = *ξ*, and *λ*(*t*) = *λ*. Thus, the SDE for *β*(*t*) now reads

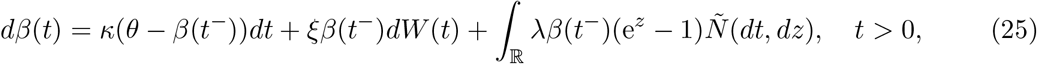

with *β*(0) = *β*_0_ known. It is worth mentioning that, again, to analyze the asymptotic properties of the transmission parameter, we must assume that its coefficients are constant.

Since *Ñ* is a compensated Poisson random measure, it follows that [49]

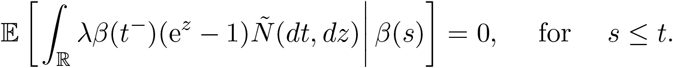

Then, *β*(*t*) in Eq. (25) also satisfies the conditional expected value in Eq. (14). This means that the limit in Eq. (16) also holds for the present process.

Using again Fubini’s theorem, the Itô’s formula for jump-diffusions [52], and the rule for the expected values of the integral with respect to the compensated Poisson measure [49], it follows that

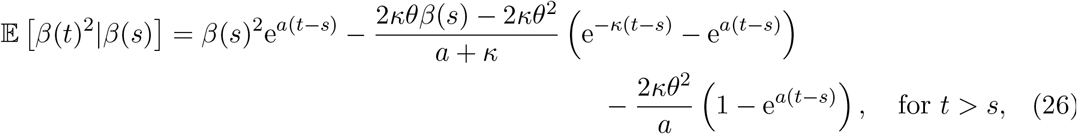

where *a* = *ξ*^2^ + *λ*^2^ ∫ _ℝ_ (e^*z*^ − 1)^2^*ν*(*dz*) − 2*κ*.

Here, we replace the conditions in Eq. (18) in Section 3.1 by

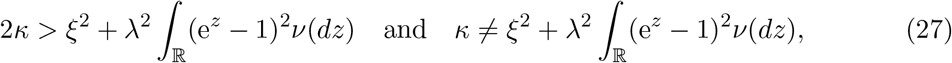

respectively. Under the conditions in Eq. (27), the quantity *a* becomes negative and it follows that

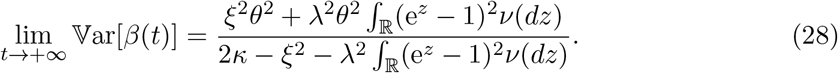

Notice that, by setting *λ* = 0, we recover the estimates in Eqs. (15)–(17) in Section 3.1 from the estimates in Eqs. (26)–(28).

#### Proposition 2.

*If the conditions in* *Eq*. (27) *are satisfied, then, the process in* *Eq*. (25) *admits at least one invariant measure*.

*Proof*. By Theorem 4.5 in [8], since the coefficients in the SDE in Eq. (25) are linear with respect to *β*_*t*_, it is only necessary to show that there are constants 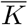 and *M*, such that

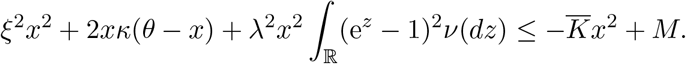

By the first condition in Eq. (27), it is only necessary to choose 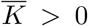 such that 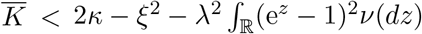 and *M* ≥ 0, such that

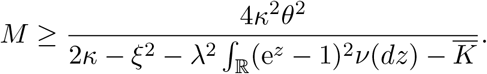

□

Sufficient conditions for the uniqueness of the invariant measure and convergence rates can be found, for example, in Corollary 5.2 in [9]. However, the processes considered so far, do not satisfy them.

By applying the Itô’s formula for jump-diffusion processes [52], it is easy to see that the infinitesimal generator associated with the SDE in Eq. (25) is the following:

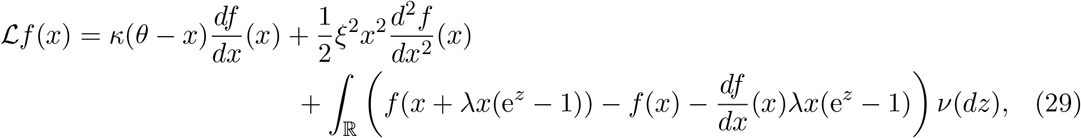

where *f* is any sufficiently regular function defined in ℝ_+_. Using integration by parts and change of variables *y* = *x* + *λx*(e^*z*^ − 1) [37], such that

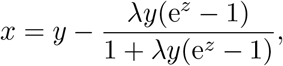

it follows that the adjoint in *L*^2^(ℝ_+_) of the operator ℒ is the following

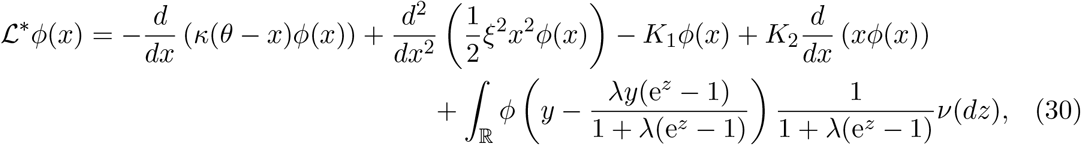

where *K*_1_ = *ν* (ℝ) and *K*_2_ = *λ* ∫ _ℝ_ (e^*z*^ − 1)*ν*(*dz*), if we assume that *ν* is a finite measure. The function *ϕ* is sufficiently regular and defined in ℝ_+_.

The steady-state version of the forward Kolmogorov equation associated to *β*(*t*) in Eq. (25) is then

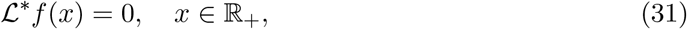

with homogeneous boundary conditions. The solution of the ODE problem in Eq. (31) defines an invariant distribution for the stochastic process *β*(*t*).

#### Remark 3.

*Under appropriate conditions, to show that μ solves the integro-differential equation in* *Eq*. (31) *in the sense of distributions, just note that*

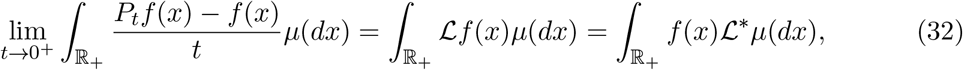

*since μ is an invariant measure, it follows that*

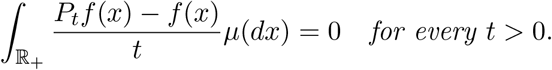

*The first equality in* *Eq*. (32) *follows by the dominated convergence theorem. It is sufficient to assume that f and its derivatives have compact support and ν is a finite measure*.

The main difficulty in finding regularity properties for a solution of the integro-differential equation in Eq. (31) is the following, it mixes degenerated coefficients in the differential part, that vanish and is unbounded, with a non-local term, i.e., the integral part. We refer the interested reader to [12, 25] that study similar problems in the context of viscosity solutions.

#### Example 2.

*The goal of this example is to evaluate the impact of the jumps and the mean-reverting speed on the probability of reaching an endemic equilibrium* ℛ _∞_ ≥ 1. *As in the previous example*, ℙ (ℛ _∞_ ≥ 1) = ℙ (*β*_∞_ ≥ 0.155). *We evaluate againg the probability of reaching the EEP, but for the model in* *Eq*. (25). *We set θ* = 0.2 *and ξ* = 0.4. *We assume that the distribution ν is Gaussian, and we test different combinations of values for its mean and standard deviation. The epidemiological-related parameters assume the values μ* = 0, *γ*_∞_ = 1*/*12 *days*^−1^, *and δ*_∞_ = 1*/*14 *days*^−1^. *The parameter κ ranges from* 0.01 *to* 2, *λ assumes the values* 0.01, 0.1, 0.5, *and* 1.0. *The mean of ν assumes the values* −1.0, 0, *and* 1.0, *whereas its standard deviation assumes the values* 0.5, 1.0, *and* 1.5.

*As λ increases, the probability of* ℛ_∞_ *to be larger than one becomes smaller. The probability increases as a function of κ, in general. If the mean of ν is* −1.0, *the probability values are less sensitive to changes in the standard deviation of ν. As the mean increases, the sensitivity w*.*r*.*t the standard deviation increases. If λ assumes the values* 0.5 *and* 1.0, *the probability becomes closer to zero, especially if the mean of ν is* 0 *or* 1.0. *It is worth mentioning that, for the sake of completeness, we are considering values that violate the first condition in* *Eq*. (27).

## 4 Numerical Results

### Transmission Parameter Estimation

The aim of this section is to illustrate the prediction capability of the models presented so far using daily reports of COVID-19 infections from New York City (NYC) [50] during the omicron outbreak at the end of 2021. To test the sensitivity of the epidemiological model in Eqs. (2)–(5) with respect to the estimated *β*, we use bootstrapping with 200 samples. Figure 3 shows the median and the corresponding 90% confidence interval (90% CI) of the estimated transmission parameter *β*(*t*) during two COVID-19 outbreaks in NYC, namely, the second outbreak at the end of 2020 and the omicron variant outbreak at the end of 2021.

**Figure 2:**
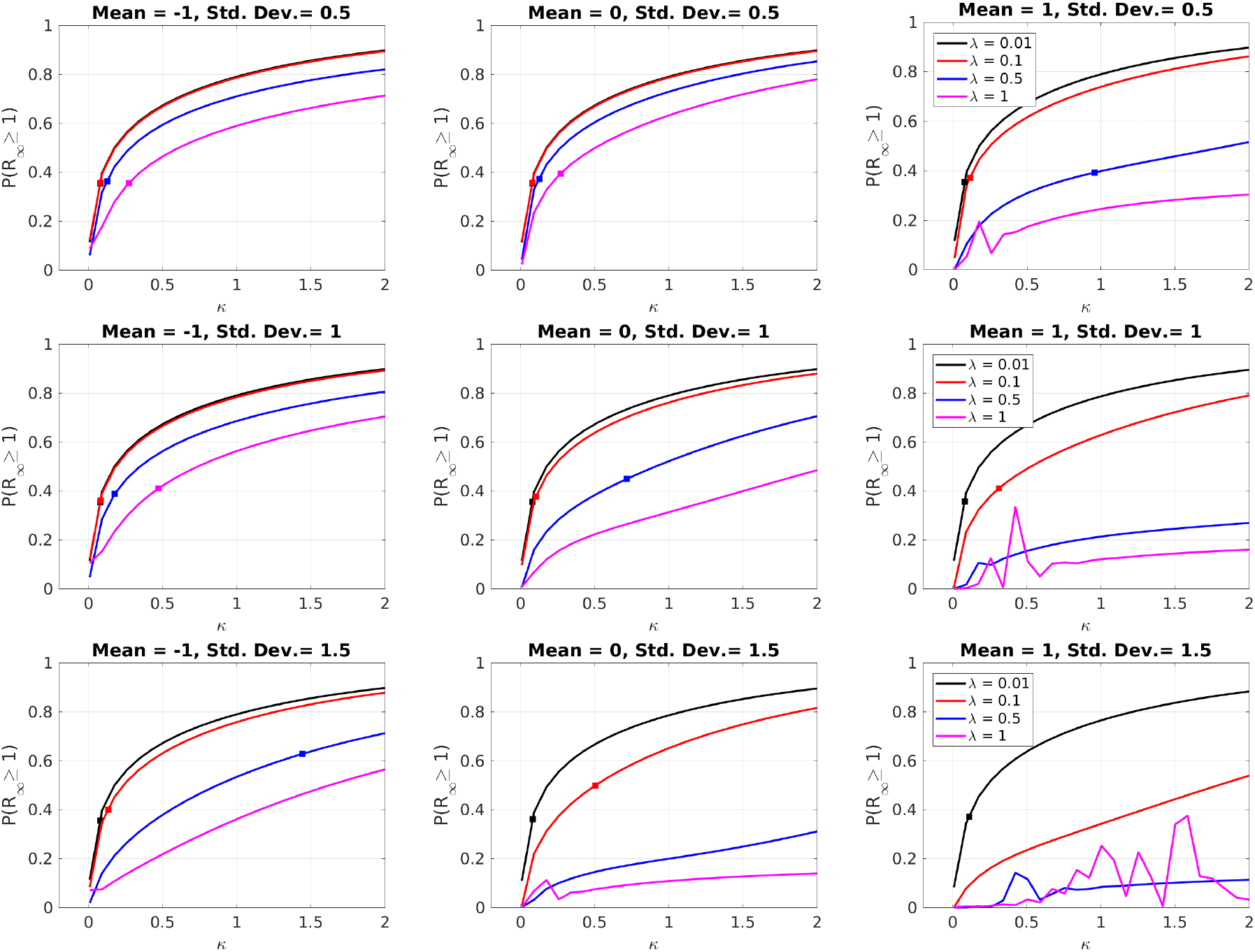
Probability of ℛ _∞_ ≥ 1 with different values for *κ, θ*, and *ξ*. The squares in the curves indicate when *κ* start to satisfy the condition in Eq. (27). If the curve does not have a square, the condition is never satisfied. The probability values for *κ* not satisfying the condition in Eq. (27) are included for completeness.

**Figure 3:**
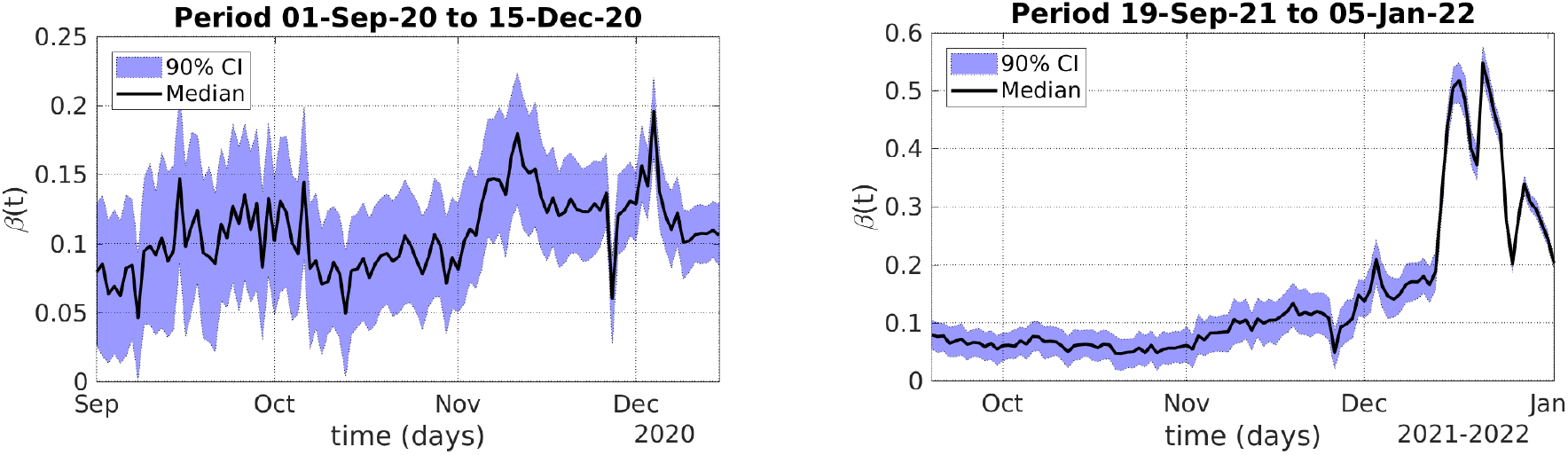
Median values and 90% CI of the estimated transmission parameter during the second COVID outbreak in 2020 (left) and the omicron outbreak at the end of 2021 (right), both in NYC. In both cases, the estimated parameter *β*(*t*) is highly volatile. During the omicron outbreak, *β*(*t*) presents a large jump occurring in the middle of December 2021.

As Figure 3 illustrates, the estimated *β*(*t*) is highly volatile. When a new and highly transmissible variant becomes dominant, as, during the omicron outbreak at the end of 2021 in NYC, we can observe jumps in the *β*(*t*) dynamics. Such patterns cannot be appropriately described by deterministic models, as many shocks in the transmission dynamics, such as the introduction of new variants, occur randomly and have an unpredictable impact. Thus, using diffusion models or, more generally, jump-diffusion models, seem to be a proper way to treat such randomness in order to improve forecasting.

It is worth mentioning that, it is important to recalibrate the models as time goes by since regime changes in a longer time horizon can cause a loss of accuracy in predictions. This is why in Eq. (6), we assume that *β*(*t*) has time-dependent parameters.

### Estimation of the Stochastic Models

After estimating the values of *β*(*t*) from the daily reported infections, we calibrate the parameters of the stochastic models from the 200 samples generated by bootstrapping. The out-of-sample predictions of the stochastic models are generated considering 5000 sample paths for each set of estimated parameters. This leads to 10^6^ sample paths. Recall that the mean-reverting (MR) model is defined in Eq. (24) and its version with jumps, which we call mean-reverting with jumps (MRJ), is defined in Eq. (33). These models have a nonlinear term in the diffusion part. The models with the linear diffusion term are the linear mean-reverting model (LMR), which is defined in Eq. (12) and its version with jumps, which we call linear mean-reverting with jumps (LMRJ), is defined in Eq. (25). For the MR and LMR models, we estimate *κ, θ*, and *ξ* from 45-day long series of *β*(*t*) values, whereas the MRJ and LMRJ models, *κ, θ, ξ, λ*, 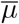, and 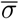 are estimated from 90-day long series to capture the jump-size distribution.

To illustrate the performance of the estimated model, in-sample and out-of-sample predictions for *β*(*t*) and for accumulated infections are provided and compared with observed data, in the case of infections. The out-of-sample predictions provide 60-day-long forecasted scenarios. To stress-test, such predictions are performed during two major outbreaks of COVID-19 in NYC, namely, the second wave of infections at the end of 2020 and the omicron variant outbreak at the end of 2021. The prediction plots can be found in Figures 4–7.

**Figure 4:**
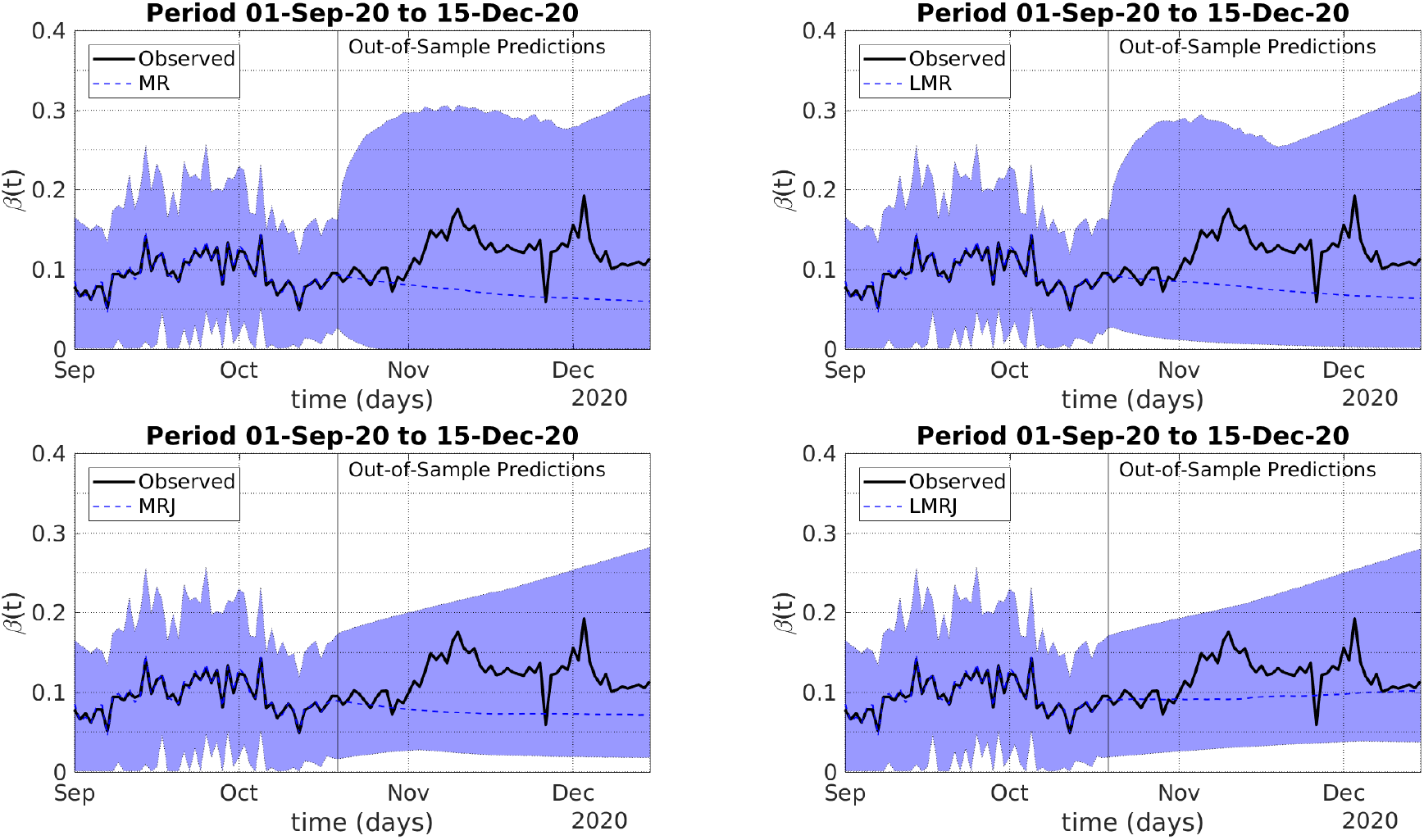
In-sample and out-of-sample model predictions for the parameter *β* during the second wave of COVID-19 infections in NYC during 2020. The out-of-sample predictions are on the right-hand side of the vertical lines. The filled envelopes represent the 90% CIs.

**Figure 5:**
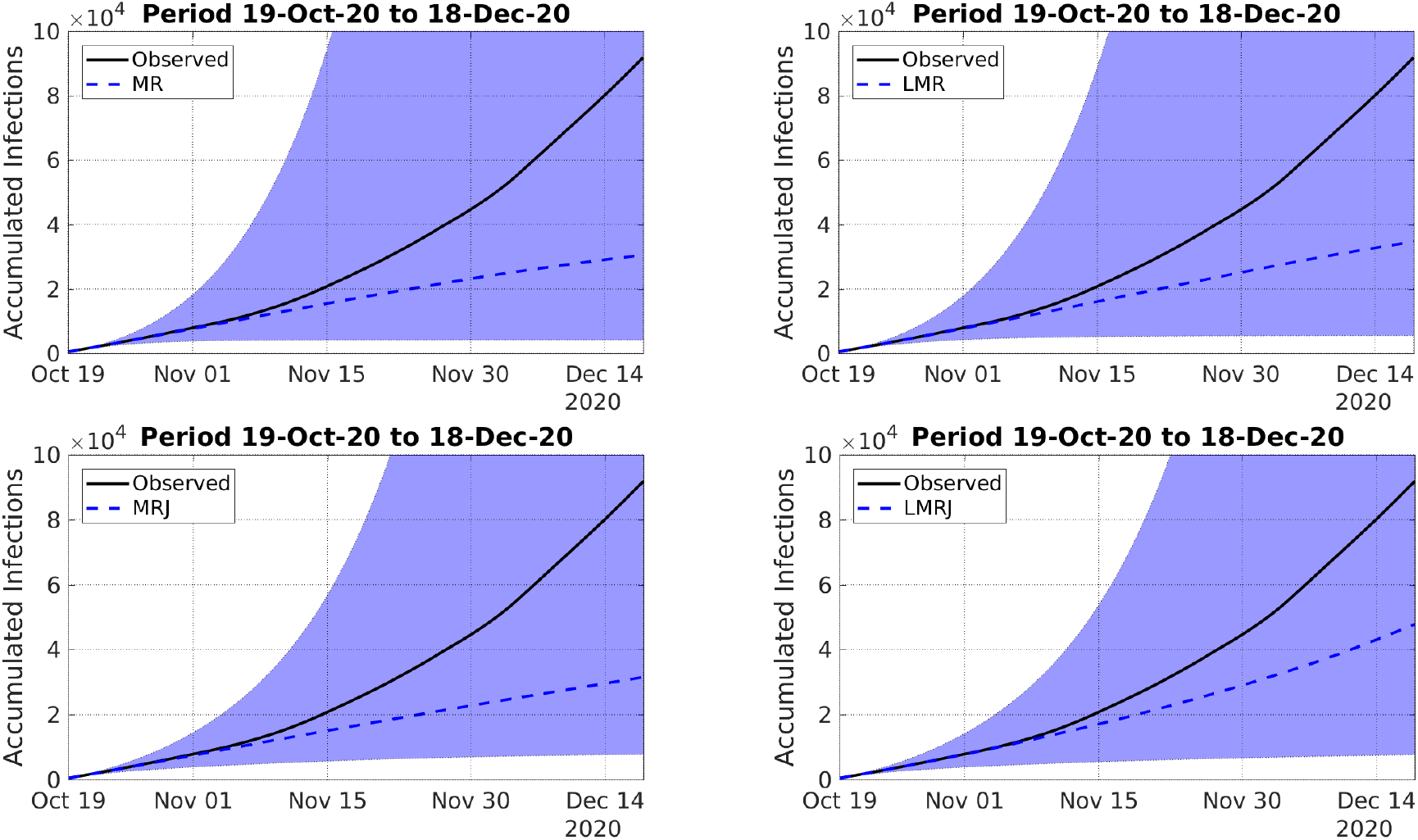
Out-of-sample model predictions for the accumulated number of infections during the second wave of COVID-19 in NYC during 2020. The filled envelopes represent the 90% CIs.

**Figure 6:**
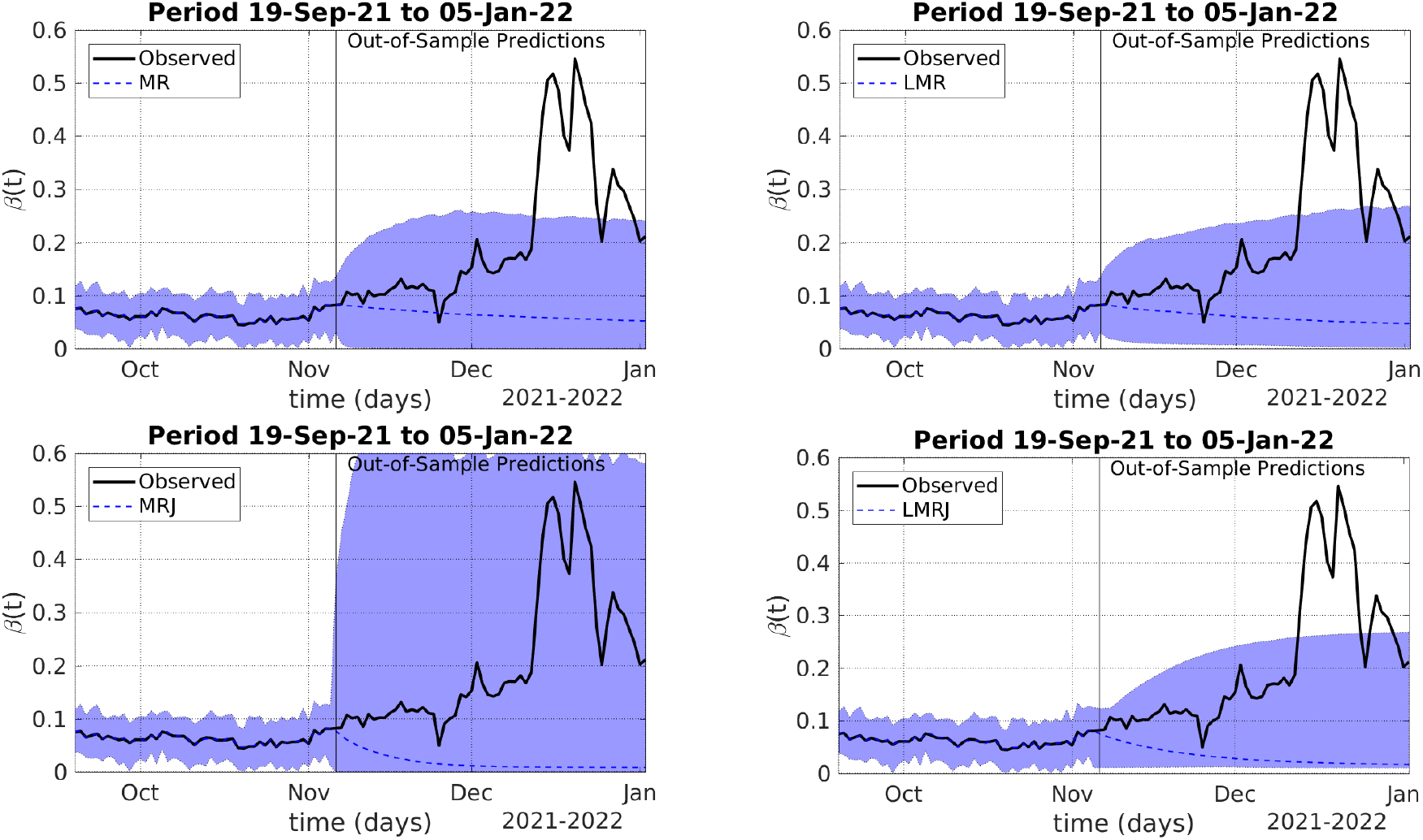
In-sample and out-of-sample model predictions for the parameter *β* during the omicron variant outbreak in NYC at the end of 2021. The out-of-sample predictions are on the right-hand side of the vertical lines. The filled envelopes represent the 90% CIs.

**Figure 7:**
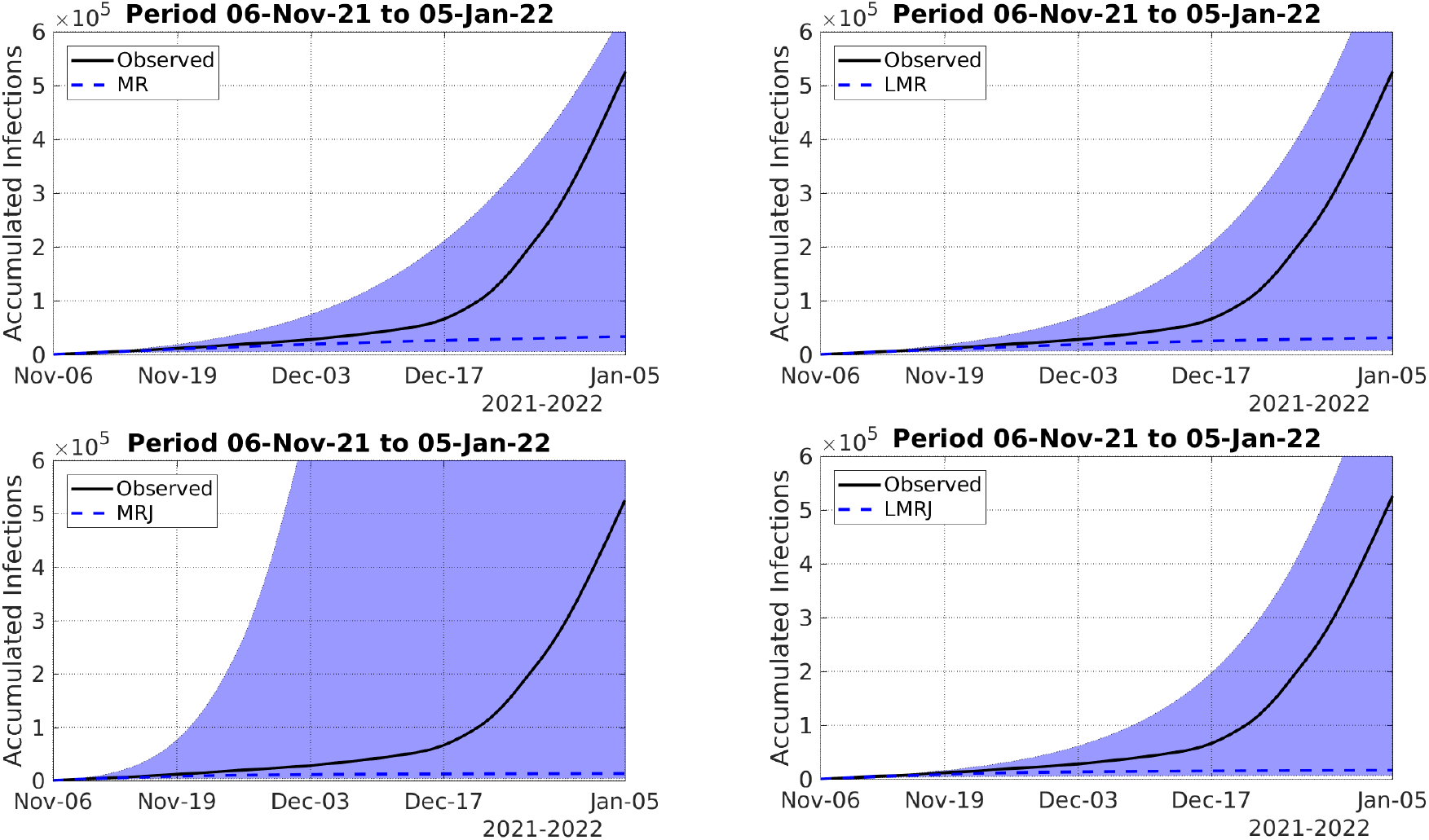
Out-of-sample model predictions for the accumulated number of infections during the omicron variant outbreak in NYC at the end of 2021. The filled envelopes represent the 90% CIs.

The estimated median and 90% CI values of the model parameters for the two NYC COVID-19 outbreaks can be found in Table 1.

**Table 1:** Median values estimated parameters for the stochastic models using data just before the NYC second outbreak of COVID-19 at the end of 2020 and just before the NYC omicron variant outbreak at the end of 2021. The numbers inside the parentheses are 90% CIs.

| Model | Parm. | 2nd. Wave (2020) | Omicron Wave (2021) |
| --- | --- | --- | --- |
| MR | $\kappa$ | 8.34 (0.58–9.69) | 7.99 (0.62–9.82) |
| | $\theta$ | 0.05 (0.00–0.48) | 0.05 (0.02–0.06) |
| | $\xi$ | 0.29 (0.10–1.02) | 0.44 (0.21–1.2) |
| LMR | $\kappa$ | 5.49 (0.27–8.14) | 5.70 (0.63–10.0) |
| | $\theta$ | 0.05 (0.00–1.40) | 0.04 (0.01–0.05) |
| | $\xi$ | 1.05 (0.10–2.39) | 1.61 (0.29–3.56) |
| MRJ | $\kappa$ | 16.6 (0.27–33.1) | 15.4 (4.24–22.8) |
| | $\theta$ | 0.07 (0.04–1.11) | 0.02 (0.01–0.03) |
| | $\xi$ | 0.36 (0.10–0.62) | 1.78 (0.18–4.65) |
| | $\lambda$ | 0.65 (0.46–5.49) | 0.68 (0.09–4.06) |
| | $\bar{\mu}$ | -2.12 (-6.78–1.46) | -0.51 (-4.7–0.72) |
| | $\bar{\sigma}$ | 1.03 (0.68–1.98) | 1.16 (0.07–1.61) |
| LMRJ | $\kappa$ | 1.86 (0.08–31.01) | 19.0 (8.07–34.4) |
| | $\theta$ | 0.24 (0.05–2.39) | 0.01 (0.01–0.20) |
| | $\xi$ | 0.30 (0.10–0.55) | 0.10 (0.10–0.50) |
| | $\lambda$ | 1.88 (0.46–5.96) | 0.70 (0.62–0.78) |
| | $\bar{\mu}$ | -1.95 (-7.19–1.24) | 0.58 (-0.43–0.91) |
| | $\bar{\sigma}$ | 1.10 (0.65–2.07) | 1.25 (0.87–1.32) |

In Figures 4 and 6, the solid black line represents the median *β*(*t*) values directly estimated from reported infections. We call such median values as observed since they are not given by any stochastic model and we use the as our benchmark for the forecasted scenarios of the transmission parameter evolution given by the stochastic models.

For the second outbreak of 2020, Figure 4 shows that the 90% CIs of the out-of-sample predictions of all four models contain the path of *β* (black solid line). However, the jump-diffusion models, i.e., MRJ and LMRJ, presented tighter 90% CIs in comparison to MR and LMR. This is reflected in the evolution of the predicted accumulated infections in Figure 5, where, the MRJ and LMRJ models also presented tighter 90% CIs containing the reported infections.

In other words, the MRJ and LMRJ models successfully incorporated the uncertainty in the data presenting more accurate results. The wider 90% CIs of the MR and LMR models is due to the larger estimated values for the diffusion parameter *ξ* shown in Table 1. As Figures 3–4 show, *β*(*t*) is highly volatile, presenting large changes during short periods. To incorporate such shocks, the MR and LMR models need artificially larger diffusion parameters. Therefore, the MRJ and LMRJ models performed better, presenting more accurate (tighter 90% CIs) results.

It is worth mentioning that, during the 2020 outbreak, all the models presented large estimated values for the mean-reverting speed (*κ*) and small estimated values for the mean *θ*. Concerning the jump-size distribution, the estimated mean values for the MRJ and LMRJ models were mainly negative, and the estimated standard deviations had the same order of magnitude as the estimated mean values. In other words, the jumps were mainly negative during this period. Of course, positive jumps could occur, as the distribution is defined in ℛ. The MRJ and LMRJ models presented similar performance and similar estimated values. However, the parameters *θ* and *λ* presented considerably larger estimated values for the LMRJ than for the MRJ. So, in the LMRJ the jump part has a larger weight in the dynamics and the long-term mean is larger. Such differences probably led the LMRJ to present a slightly tighter 90% CI for the *β*(*t*) predictions, with median values closer to observed *β*(*t*) values.

As Figures 6–7 show, during the NYC omicron variant outbreak, at the end of 2021, MR, LMR, and LMRJ performed quite similarly, presenting relatively tight 90% CIs for the out-of-sample predictions of *β*(*t*), and, in consequence, for the accumulated infections. None of these three models was able to capture the larger jumps in the *β*(*t*) time evolution inside their 90% CIs, however, accumulated infection predictions were accurate. The MRJ presented a much wider 90% CI for out-of-sample predictions for *β*(*t*), which contained the observed large jumps. However, the 90% CI of accumulated infections predictions was considerably large, which may lead to unrealistic scenarios. Such a wider 90% CI for MRJ is linked to the large values for the estimated diffusion parameter *ξ*, in comparison to MR and LMRJ. The LMRJ presented mainly positive jumps, with the jump part having a smaller role in the dynamics than in the 2020 outbreak.

### Long-Term Predictions

To illustrate the performance of long-term predictions, we provide a 340-day long forecast, corresponding to the period from 11-Oct-2021 to 15-Sept-2022. Figures 9–8 present the forecasted daily infections and beta values.

**Figure 8:**
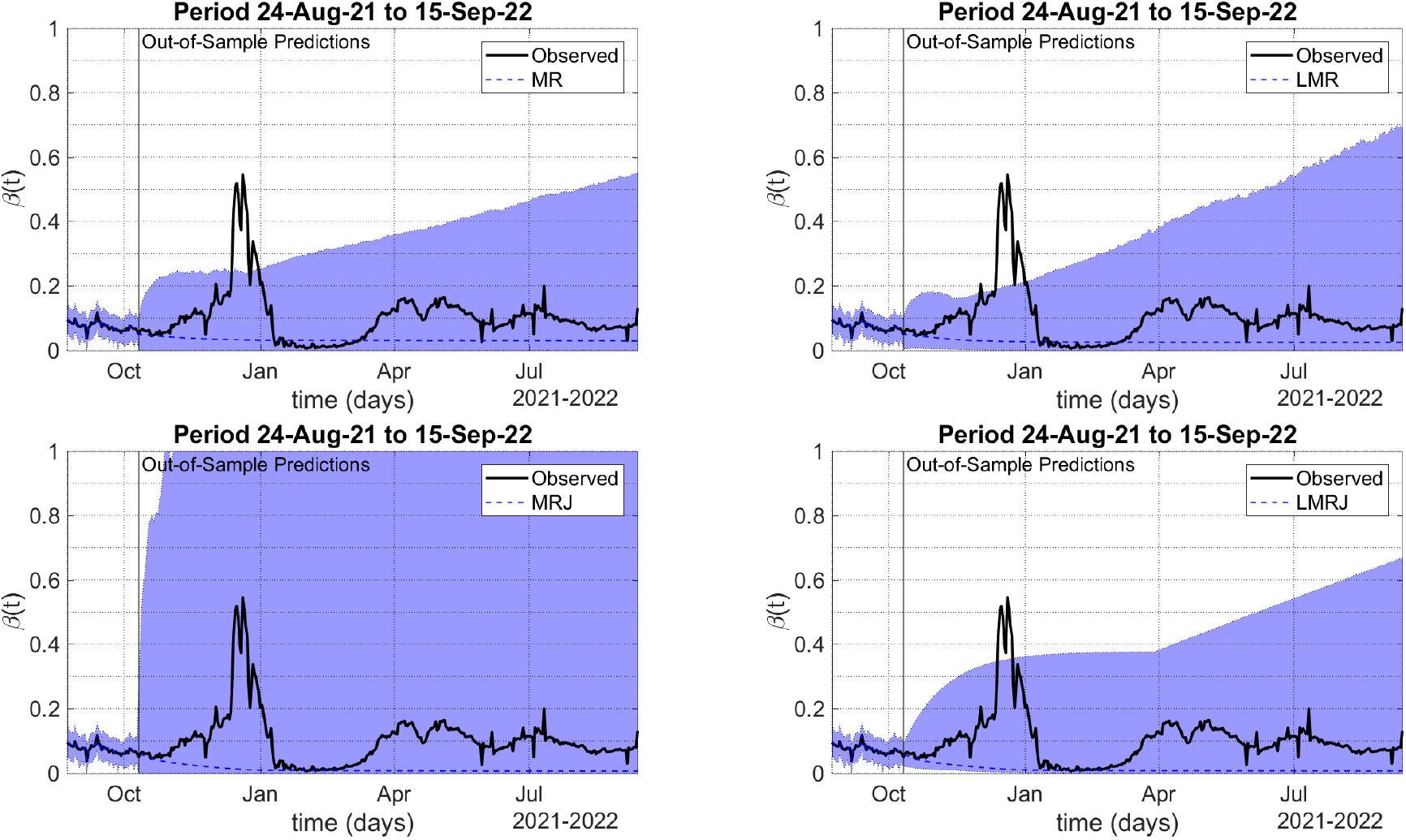
In-sample and out-of-sample model predictions for the parameter *β* during the omicron variant outbreak in NYC at the end of 2021. The out-of-sample predictions are on the right-hand side of the vertical lines. The filled envelopes represent the 90% CIs.

**Figure 9:**
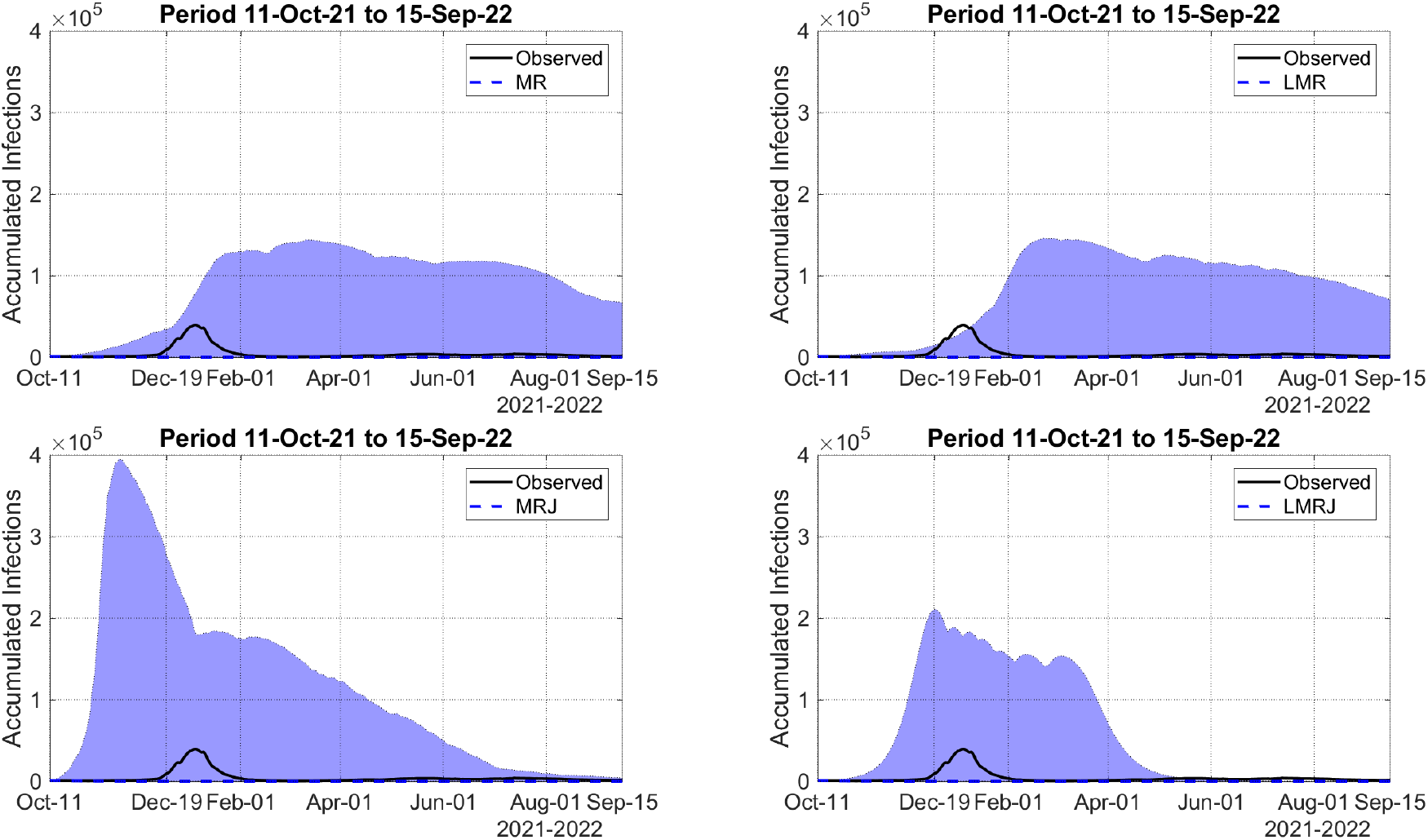
Out-of-sample model predictions for the daily number of infections. The filled envelopes represent the 90% CIs.

As Figure 8 show, the MRJ model presented a much wider 90% CI for *β*(*t*) predictions, leading to a massive outbreak in Nov-Dec 2021, shown in Figure 9, anticipating the actual omicron outbreak. After that, apparently, the disease dies out. The MR and LMR models had very similar performances, with similar 90% CIs for *β*(*t*) and infections predictions. In both cases, the model predicts long-standing outbreaks during 2022. However, in the LMR predictions, the outbreak starts just after the beginning of the actual omicron outbreak. The LMRJ model also predicts a large outbreak, containing the actual omicron outbreak.

However, it is smaller than the outbreak predicted by the MRJ model and lasts for a smaller period in comparison to the other model predictions. In other words, once again, the LMRJ seems to outperform the other models.

### Asymptotic Behavior

We now use the estimated parameter values of the LMR and LMRJ models corresponding to Figures 8–9 to evaluate the probability of the effective reproduction number ℛ (*t*) be larger than one, when *t* → ∞. The procedure is the same performed in Examples 1–2. The probability values are evaluated considering the estimated parameters that satisfy the conditions in Eqs. 18 and (27). Such parameters represent 89,0% and 73,5% of the estimated values of the LMR and LMRJ, respectively.

For the LMR, the probability has a median value of 0.74% (70% CI: 0%–2.08%), and for the LMRJ, it has a median value of 0,04% (70% CI: 0,01%–35,18%). Although the LMRJ has a smaller median value, the 70% CI is much wider. This means that reaching a disease-free equilibrium, with a transmission process driven by a jump-diffusion model is unlikely, considering the estimation period, i.e., from 13-Jul to 10-Oct-21. This is in agreement with the observed data, that show recurrent outbreaks in the period of forecast. Moreover, the median value of the best-fit estimation of ℛ (*t*) during the period 11-Oct-21 to 15-Sept-22 is 0.58 (70% CI: 0.17–0.94), with 12,7% of its values larger than one.

### Long-term Predictions with Immunity Loss

As an additional experiment, we add the possibility of loss of immunity for those recovered. Thus, we add to Eq. (4) the term −*αR*, and to Eq. (2), we add +*αR*. In this example, *α* assumes the value 1*/*360 days^−1^. The forecasted period spans over 3 years, starting at 11-Oct-2021, which contains the omicron variant outbreak. Figure 10 presents the model predictions.

**Figure 10:**
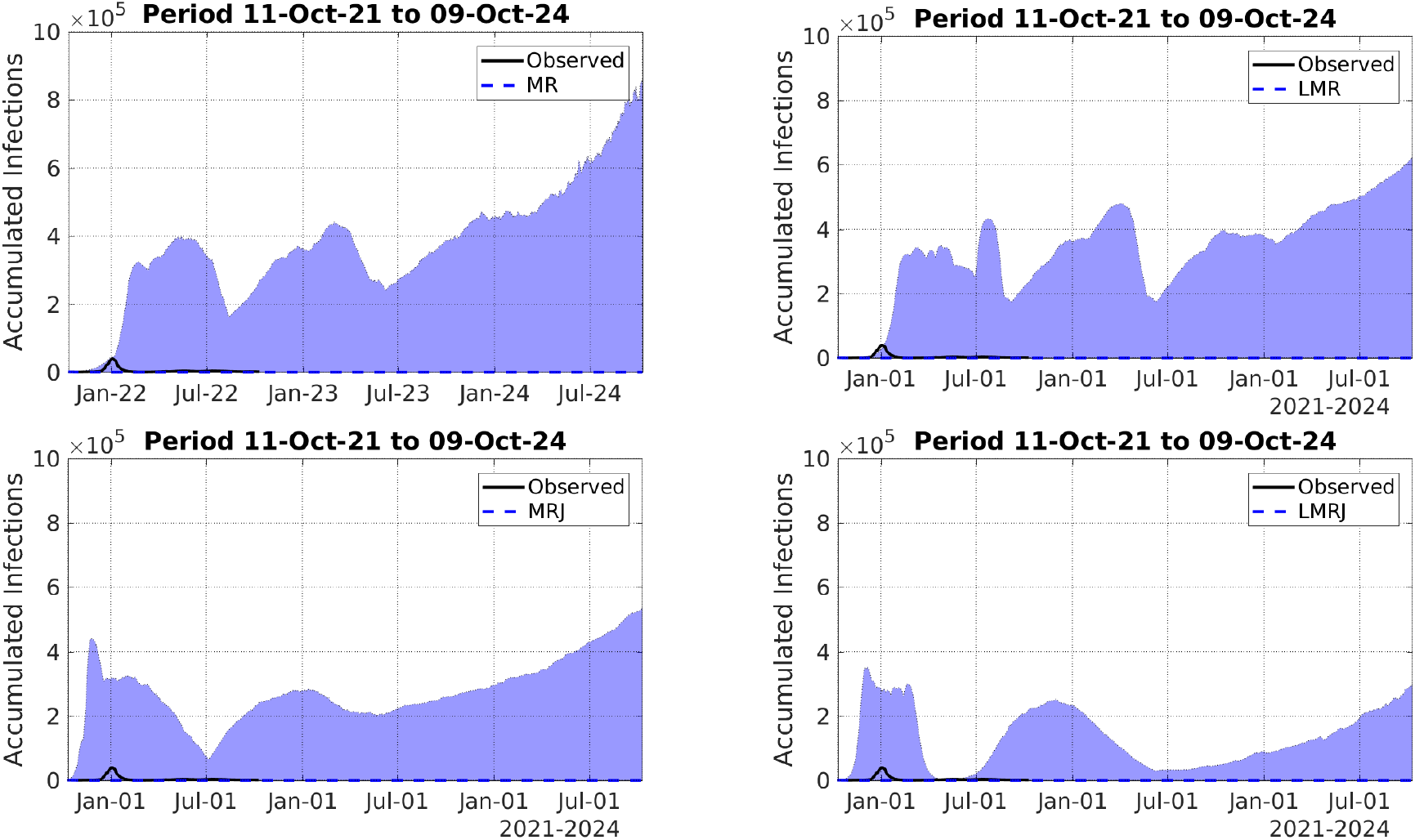
Out-of-sample model predictions for the daily number of infections considering the loss of immunity. The filled envelopes represent the 90% CIs.

Again, the MRJ and LMRJ models presented more accurate predictions, with less massive waves of infections. Moreover, the predictions provided by the LMRJ seem more plausible. Both models presented recurrent waves when the loss of immunity occurs with a mean time of 360 days. The other models also presented recurrent waves, however, with magnitudes much higher than the omicron outbreak.

## 5 Discussion and Concluding Remarks

Quantitative Finance (QF) teaches us that we must identify the main sources of uncertainties in the market to create parsimonious models that properly incorporate the data variability and stylized facts to provide accurate forecasts [43]. By using stochastic tools in epidemiological models, we take advantage of the accumulated wisdom from QF, building parsimonious and computationally efficient models, which can be tested with real data thus producing forecasts that can be compared with actual reported data. By comparing out-of-sample model predictions with observed data, in different situations, it is possible to evaluate the model performance.

In the present work, we followed these steps, building stochastic adaptations of the classical SEIR model [41], focusing on modeling the time evolution of the transmission parameter *β*. We do not add stochastic terms to the SEIR compartments’ dynamics for a number of reasons. Firstly, because *β* depends on the average number of daily contacts and the probability of such contacts will result in infection, these two components, the average number of contacts and the infection probability, are time-dependent random values. So, it seems natural to assume that *β* is a stochastic process. Secondly, adding stochastic terms to the other equations in the model may not improve its performance, since it will necessarily increase the model complexity and the stochastic components may have some sort of correlation that must be estimated from data. In addition, the data may be not sufficient to calibrate the additional number of parameters.

The chosen structure for *β*(*t*) is in line with the estimations. After observed shocks, which can lead to large values during outbreaks or smaller values during lockdowns and other contention measures, *β*(*t*) tends to return to some mean level. This justifies the use of a mean-reverting term. Indeed, periods of high incidence lead to contention measures, and periods of low incidence lead to relaxation of the contention measures. Moreover, the observed fluctuations in the estimated values at any time can be modeled as diffusion. The jump term adds an extra degree of freedom that accommodates sudden shocks in the dynamics that cannot be well parameterized by a diffusion term, as observed during larger outbreaks in NYC, such as the second wave in 2020 and the omicron outbreak that started at the end of 2021.

It is well-known that small jumps can be viewed as diffusion [24]. Thus, during periods when transmission stabilizes, diffusion models and jump-diffusion models will perform similarly. However, during major outbreaks, jump-diffusion models can address better the uncertainty in the data providing potentially more accurate predictions. This is illustrated by the results obtained using the LMRJ, which presented accurate out-of-sample predictions during two major outbreaks in NYC, outperforming the other models.

For the sake of simplicity, in this study, we did not consider a series of features observed in the COVID-19 dynamics, such as differences in the degree of the disease severity driven by age and sex, the effects of vaccination, loss of immunity and reinfection, asymptomatic infection, and underreporting. Some of these features were already studied in previous works [6, 5, 7]. Immunity loss must be one of the main reasons for the emergence of new outbreaks. Furthermore, data on reinfection and infections after vaccination is absent or, at least, difficult to access. Thus, calling for a modelling approach to immunity loss. In future work, we aim to investigate the relationship between the loss of immunity and jumps in the transmission dynamics.

## Data Availability

All data produced in the present work are contained in the manuscript.

https://github.com/viniciusalbani/StochasticTransmission

## A Appendix

### A.1 Numerical Implementation

In this section, we present numerical tools to simulate and calibrate the stochastic models presented so far to make out-of-sample predictions of infections. All the codes used in this work are available from the GitHub repository https://github.com/viniciusalbani/StochasticTransmission.

In this example, the performance of four stochastic models are compared, namely, the CIR model in Eq. (24) and its version with jumps,

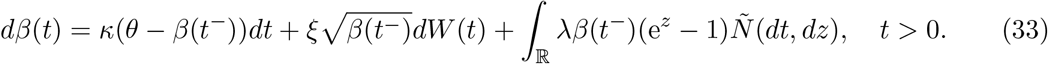

as well as the models in Eq.(12) and Eq. 25.

#### Calibration Procedure

The model calibration is divided into the following two steps:

1. Estimate the values of *β*(*t*) from daily reports of infections by minimizing the functional

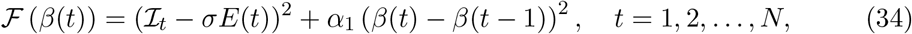

with *β*(0) given and *α*_1_ *>* 0 the regularization parameter.
2. As in [3], the parameters of the models describing *β*(*t*) are estimated from the values obtained in the first step. This procedure helps to improve model adherence to the data.

To implement Step 2 above, as *β*(*t*) is observed in discrete time, we minimize the following functional:

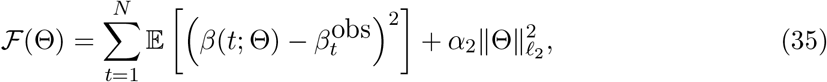

where Θ represents the vector of the parameters defining the model for *β*(*t*), 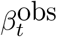 denotes the value for *β*(*t*) estimated in Step 1 above, and *α*_2_ is the regularization parameter. Notice that

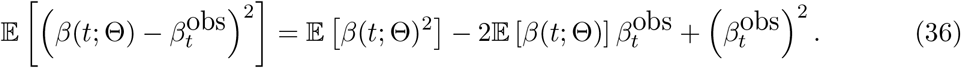

To evaluate the expected values in Eq. (36), we apply the Itô formula, discretize the SDEs for *β*(*t*) considering an Euler-Maruyama-like scheme [38], and take expectations finding for the CIR model with jumps,

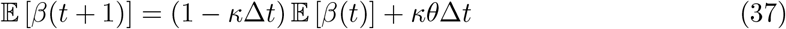

and

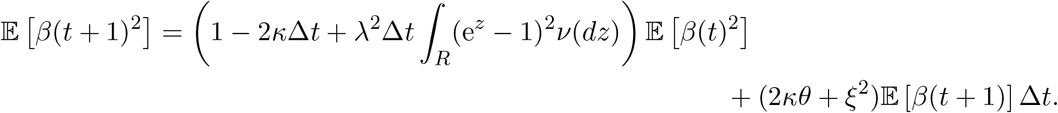

For the model in Eq. (25), the expected value 𝔼 [*β*(*t* + 1)] is equal to the one in Eq. (37), whereas

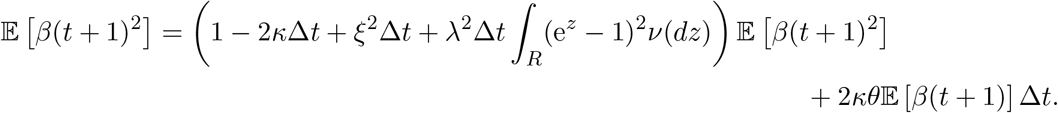

To evaluate the expected values above for the model versions without jumps, just set *λ* = 0.

In this numerical example, we further assume that *λ* = 1 in the cases with jumps and the jump-size distribution *ν* is Gaussian with mean *μ* and variance *σ*^2^, i.e.,

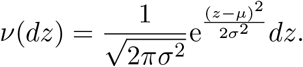

Thus, 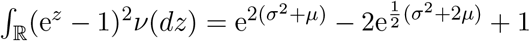.

#### Model Simulation

To generate scenarios using the models with jumps in Eqs. (25) and (33), we use a numerical scheme for jump-diffusion models proposed in [33]. The versions without jumps are solved numerically by the Euler-Maruyama scheme [38].

The minimization of the objective functions in Eqs. (34)–(35) is performed by using the 2022 release of MATLAB’s routine LSQNONLIN.

#### Evaluation of Example 2

To calculate ℙ (*β*_∞_ ≥ 0.155) in Example 2, we evaluate the integral

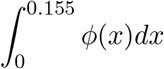

by the trapezoidal rule. To evaluate *ϕ*, we numerically solve the integro-differential equation in Eq. (31) by a finite-difference scheme for the differential part and the trapezoidal rule for the integral part. Then we build a fixed-point iteration to approximate *ϕ*, where, at each iteration, a linear system is solved with the integral part approximation on the right-hand side. More precisely, if *ϕ*^0^ represents the initial step, we solve the iterations

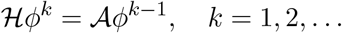

We repeat the iterations until 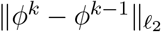 is sufficiently small.

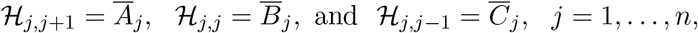

where *n* is the number of discretization points, Δ*x* is the step-size, 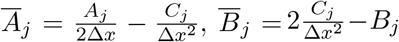, and 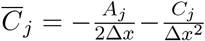, with 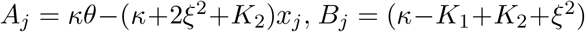, and 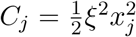. In Example 2, *K*_1_= 1, and 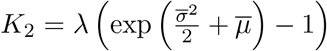, where 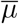 and 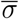 denote the mean and variance of the Gaussian distribution *ν*. The operator 𝒜 is the trapezoidal rule applied to the discrete version of the function

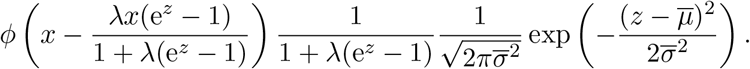

## References

[1] M. Achterberg, B. Prasse, L. Ma, S. Trajanovski, M. Kitsak, and P. Van Mieghem, Comparing the accuracy of several network-based COVID-19 prediction algorithms, International Journal of Forecasting (2020).

[2] V. Albani, R. Albani, N. Bobko, E. Massad, and J. Zubelli, On the role of financial support programs in mitigating the SARS-CoV-2 spread in Brazil, BMC Public Health 22 (2022), no. 1, 1–17.

[3] V. Albani, R. Albani, E. Massad, and J. Zubelli, Nowcasting and Forecasting COVID-19 Waves: The Recursive and Stochastic Nature of Transmission, Royal Society Open Science 9 (2022), 220489.

[4] V. Albani, M. Grasselli, W. Peng, and J. Zubelli, The interplay between covid-19 and the economy in canada, Journal of Risk and Financial Management 15 (2022), no. 10.

[5] V. Albani, J. Loria, E. Massad, and J. Zubelli, COVID-19 Underreporting and its Impact on Vaccination Strategies, BMC Infectious Diseases 21 (2021), 1111.

[6] V. Albani, J. Loria, E. Massad, and J.P. Zubelli, The Impact of COVID-19 Vaccination Delay: A Data-Driven Modelling Analysis for Chicago and New York City, Vaccine 39 (2021), no. 41, 6088–6094.

[7] V. Albani, R. Velho, and J. Zubelli, Estimating, Monitoring, and Forecasting the Covid-19 Epidemics: A Spatio-Temporal Approach Applied to NYC Data, Scientific Reports (2021), 9089.

[8] S. Albeverio, Z. Brzeźniak, and J.-L. Wu, Existence of global solutions and invariant measures for stochastic differential equations driven by Poisson type noise with non-Lipschitz coefficients, Journal of Mathematical Analysis and Applications 371 (2010), no. 1, 309–322.

[9] A. Arapostathis, G. Pang, and N. Sandrić, Ergodicity of a lévy-driven sde arising from multiclass many-server queues, The Annals of Applied Probability 29 (2019), no. 2, 1070–1126.

[10] A. Aspri, E. Beretta, A. Gandolfi, and E. Wasmer, Mortality containment vs. economics opening: optimal policies in a SEIARD model, Journal of Mathematical Economics 93 (2021), 102490.

[11] G. Athayde and A. Alencar, Forecasting Covid-19 in the United Kingdom: A dynamic SIRD model, PLoS ONE 17 (2022), e0271577.

[12] G. Barles and C. Imbert, Second-order elliptic integro-differential equations: viscosity solutions’ theory revisited, Annales de l’IHP Analyse Non Linéaire 25 (2008), no. 3, 567–585.

[13] M. Bartlett, Deterministic and stochastic models for recurrent epidemics, Proc. Third Berkeley Symposium on Mathematical Statistics and Probability 4 (1956), 81–109.

[14] Some stochastic models in ecology and epidemiology, Contributions to Probability and Statistics, A Volume dedicated to Harold Hotelling (1960), 89–96.

[15] Nicola Bellomo, Richard Bingham, Mark AJ Chaplain, Giovanni Dosi, Guido Forni, Damian A Knopoff, John Lowengrub, Reidun Twarock, and Maria Enrica Virgillito, A multiscale model of virus pandemic: Heterogeneous interactive entities in a globally connected world, Mathematical Models and Methods in Applied Sciences 30 (2020), no. 08, 1591–1651.

[16] Nicola Bellomo, Diletta Burini, and Nisrine Outada, Multiscale models of Covid-19 with mutations and variants, Networks and Heterogeneous Media 17 (2022), no. 3, 293.

[17] A. Bertozzi, E. Franco, G. Mohler, M. Short, and D. Sledge, The challenges of modeling and forecasting the spread of COVID-19, Proceedings of the National Academy of Sciences 117 (2020), no. 29, 16732–16738.

[18] C. Bianca and C. Dogbe, On the existence and uniqueness of invariant measure for multidimensional stochastic processes, Nonlinear Studies-The International Journal (2017).

[19] O. Bjørnstad, B. Finkenstädt, and B. Grenfell, Dynamics of measles epidemics: estimating scaling of transmission rates using a time series SIR model, Ecological Monographs 72 (2002), no. 2, 169–184.

[20] T. Britton, Stochastic epidemic models: a survey, Mathematical Biosciences 225 (2010), no. 1, 24–35.

[21] D. Calvetti, A. Hoover, J. Rose, and E. Somersalo, Bayesian dynamical estimation of the parameters of an SE(A)IR COVID-19 spread model, 2020.

[22] Metapopulation Network Models for Understanding, Predicting, and Managing the Coronavirus Disease COVID-19, Frontiers in Physics 8 (2020), 261.

[23] E. Campos, R. Cysne, A. Madureira, and G. Mendes, Multi-generational sir modeling: Determination of parameters, epidemiological forecasting and age-dependent vaccination policies, Infectious Disease Modelling 6 (2021), 751–765.

[24] R. Cont and P. Tankov, Financial Modelling with Jump Processes, CRC Financial Mathematics Series, Chapman and Hall, 2003.

[25] R. Cont and E. Voltchkova, Integro-differential equations for option prices in exponential Lévy models, Finance Stoch 9 (2005), no. 3, 299–325.

[26] J. Cox, J. Ingersoll Jr, and S. Ross, A theory of the term structure of interest rates, Theory of Valuation, World Scientific, 2005, pp. 129–164.

[27] O. Diekmann, J. Heesterbeek, and J. Metz, On the definition and the computation of the basic reproduction ratio R_0_ in models for infectious-diseases in heterogeneous populations, Journal of Mathematical Biology 28 (1990), no. 4, 365–382.

[28] R. Engbert, M. Rabe, R. Kliegl, and S. Reich, Sequential data assimilation of the stochastic SEIR epidemic model for regional COVID-19 dynamics, Bulletin of Mathematical Biology 83 (2021), no. 1, 1–16.

[29] L.C. Evans, An Introduction to Stochastic Differential Equations, vol. 82, American Mathematical Society, 2012.

[30] D. Faranda and T. Alberti, Modeling the second wave of COVID-19 infections in France and Italy via a stochastic SEIR model, Chaos: An Interdisciplinary Journal of Nonlinear Science 30 (2020), no. 11, 111101.

[31] A. F. Filippov, Equations with the right-hand side continuous in x and discontinuous in t, pp. 3–47, Springer Netherlands, Dordrecht, 1988.

[32] M. Gatto, E. Bertuzzo, L. Mari, S. Miccoli, L. Carraro, R. Casagrandi, and A. Rinaldo, Spread and dynamics of the covid-19 epidemic in italy: Effects of emergency containment measures, Proceedings of the National Academy of Sciences 117 (2020), no. 19, 10484– 10491.

[33] K. Giesecke, G. Teng, and Y. Wei, Numerical Solution of Jump-Diffusion SDEs, SSRN Preprint, 2018.

[34] A. Gray, D. Greenhalgh, L. Hu, X. Mao, and J. Pan, A stochastic differential equation SIS epidemic model, SIAM Journal on Applied Mathematics 71 (2011), no. 3, 876–902.

[35] N. Guglielmi, E. Iacomini, and A. Viguerie, Delay differential equations for the spatially resolved simulation of epidemics with specific application to COVID-19, Mathematical Methods in the Applied Sciences (2022).

[36] M. Hairer, Convergence of Markov processes, (2021).

[37] F. B. Hanson, Applied stochastic processes and control for jump-diffusions: Modeling, analysis and computation, Society for Industrial and Applied Mathematics, Philadelphia, PA, 2007.

[38] D. Higham, An Algorithmic Introduction to Numerical Simulation of Stochastic Differential Equations, SIAM Rev. 43 (2001), no. 3, 525–546.

[39] J. Jacquez and C. Simon, The stochastic SI model with recruitment and deaths I. Comparison with the closed SIS model, Mathematical Biosciences 117 (1993), no. 1-2, 77–125.

[40] I. Karatzas and S. Shreve, Brownian Motion and Stochastic Calculus, vol. 113, Springer, 2012.

[41] M.J. Keeling and R. Rohani, Modeling infectious diseases in humans and animals, Princeton University Press, 2008.

[42] C. Kerr, R. Stuart, D. Mistry, R. Abeysuriya, K. Rosenfeld, G. Hart, R. Núñez, J. Cohen, P. Selvaraj, B. Hagedorn, et al., Covasim: an agent-based model of COVID-19 dynamics and interventions, PLOS Computational Biology 17 (2021), no. 7, e1009149.

[43] R. Korn and E. Korn, Option Price and Portfolio Optimization: Modern Methods of Mathematical Finance, Graduate Studies in Mathematics, vol. 31, AMS, 2001.

[44] A. Korobeinikov, Global properties of SIR and SEIR epidemic models with multiple parallel infectious stages, Bulletin of mathematical biology 71 (2009), no. 1, 75–83.

[45] A. Leitao and C. Vázquez, The stochastic θ-SEIHRD model: Adding randomness to the COVID-19 spread, Communications in Nonlinear Science and Numerical Simulation 115 (2022), 106731.

[46] D. Melesse and A. Gumel, Global asymptotic properties of an SEIRS model with multiple infectious stages, Journal of Mathematical Analysis and Applications 366 (2010), no. 1, 202–217.

[47] S. Namasudra, S. Dhamodharavadhani, and R. Rathipriya, Nonlinear neural network based forecasting model for predicting covid-19 cases, Neural Processing Letters (2021), 1–21.

[48] I. Nåsell, Stochastic models of some endemic infections, Mathematical Biosciences 179 (2002), no. 1, 1–19.

[49] D. Nualart and E. Nualart, Introduction to Malliavin Calculus, vol. 9, Cambridge University Press, 2018.

[50] NYC, Covid-19 data from nyc https://www1.nyc.gov/site/doh/covid/covid-19-data.page.

[51] B. Øksendal, Stochastic Differential Equations: an Introduction with Applications, Springer, 2013.

[52] B. Øksendal and A. Sulem, Stochastic Control of Jump Diffusions, Springer, 2005.

[53] Nicolas Privault and Liang Wang, Stochastic sir lévy jump model with heavy-tailed increments, Journal of Nonlinear Science 31 (2021), no. 1, 1–28.

[54] C.J. Ridler-Rowe, On a stochastic model of an epidemic, Journal of Applied Probability 4 (1967), no. 1, 19–33.

[55] R. Stewart, S. Erwin, J. Piburn, N. Nagle, J. Kaufman, A. Peluso, J. Christian, J. Grant, A. Sorokine, and B. Bhaduri, Near real time monitoring and forecasting for COVID-19 situational awareness, Applied Geography 146 (2022), 102759.

[56] P Whittle, The outcome of a stochastic epidemic—a note on Bailey’s paper, Biometrika 42 (1955), no. 1-2, 116–122.

[57] Xianghua Zhang and Ke Wang, Stochastic seir model with jumps, Applied Mathematics and Computation 239 (2014), 133–143.

[58] L. Zhu, Limit theorems for a Cox-Ingersoll-Ross process with Hawkes jumps, Journal of Applied Probability 51 (2014), no. 3, 699–712.

